# A Natural Experiment Reveals Clinically Essential and Compliance-Driven Nursing Documentation

**DOI:** 10.64898/2026.06.23.26355993

**Authors:** Hao Fan, Rosemary Mugoya, Amy Finnegan, Jennifer Thate, Haomiao Jia, Sarah Rossetti, Po-Yin Yen

## Abstract

Despite contributing substantially to clinician burnout, nursing documentation lacks empirical evidence distinguishing clinically essential from administratively driven documentation. Exploiting a COVID-19 documentation relaxation policy as a natural experiment, we analyzed 520,357 patient shifts from 36,321 patients in 54 inpatient units (2019 – 2022) using large language model-assisted flowsheet classification and structural equation modeling. When permitted, front-line nurses reliably distinguished two types of documentation: in acute care units, primary nurses reduced compliance-driven Cares & Safety documentation by 19% (106.4 to 86.2 entries, r = −0.19), while maintaining or increasing documentation directly relevant to respiratory management, with no impact on patient respiratory outcomes. Documentation intensity also co-varied with real-time patient deterioration, consistently across unit types (|β| = 0.13 – 0.14). Together, these findings provide the first large-scale quantitative evidence distinguishing clinically essential documentation from compliance-driven documentation and demonstrate that targeted reduction of the latter is a viable strategy for alleviating documentation burden without compromising care quality for respiratory care management.

## INTRODUCTION

Inpatient nurses spend roughly one-quarter of every shift, more than two hours out of twelve, interacting with the electronic health record (EHR), much of it on documentation unrelated to direct patient care^1^. This reflects the inherently multi-purpose nature of nursing documentation, which serves to record, synthesize, and communicate patient information, thereby bridging patient-oriented bedside care and providing foundations for billing purposes, Centers for Medicare and Medicaid Services (CMS) quality metrics, and best-practice audits^2,3^. These competing demands have driven substantial growth in nursing documentation volume and complexity^1,4,5^, amplifying the time required for reviewing the patient record and data entry^6,7^. While documentation demands and EHR usability issues (e.g., cumbersome navigation, redundant data entry, and poorly aligned workflows) detract from clinicians’ completion of core clinical tasks^8–11^, the resulting strain, commonly termed excessive documentation burden (ExDocBurden)^9^, has been widely linked to clinician stress and burnout^12–18^ and has raised concerns about downstream effects on healthcare quality and patient outcomes^19–21^.

Yet efforts to reduce ExDocBurden face a fundamental obstacle: the absence of evidence distinguishing clinically essential documentation (charting that directly informs patient care) from administratively or compliance driven documentation (charting that fulfills compliance requirements but carries little clinical signal). This distinction is further confounded by patient acuity, as sicker patients require more care and generate more documentation^22–24^, making it challenging to assess the relationship between documentation practices and clinical outcomes^25,26^. Consequently, the downstream effects of documentation reduction on patient outcomes remain largely unknown.

The coronavirus disease 2019 (COVID-19) pandemic offered a unique opportunity to conduct a natural experiment, when federal- and state-level policies temporarily suspended specific administrative documentation requirements during surge periods, defined as intervals of sharply elevated COVID-19 hospitalizations that strained hospital capacity and staffing^27^. Clinicians were instructed to chart only information necessary for direct patient care guided by their clinical reasoning and judgment, which we define as *essential documentation*. In practice, hospitals operationalized these relaxations differently within their internal policies, and the present study examines one such implementation at an academic medical center in the Midwestern United States (US), hereafter referred to as the Surge Documentation policy. Notably, this policy was applied in acute care units (ACUs) but not in intensive care units (ICUs), even though both experienced the same pandemic conditions. This context enabled us to contrast *essential documentation* during the policy relaxation phases with *all-inclusive documentation* (i.e., all documentation inclusive of *essential* and *compliance-driven documentation*) before and after policy implementations^28^.

In a prior study, we demonstrated that total documentation volume decreased during the relaxation phase; however, *essential documentation* volume was not reduced^28^. What remained unknown, however, was whether those reductions affected patient outcomes. Moreover, although nursing documentation patterns are established signals of clinical surveillance predictive of patient deterioration^29^, it remained unclear how this surveillance function operates when administrative or compliance-driven documentation is relaxed. Understanding these dynamics is essential to building an evidence base that can guide documentation reform without compromising care.

In this study, we examined how nurses’ documentation patterns aligned with patient care needs in response to policy changes in documentation requirements, and whether documentation practices influenced subsequent patient respiratory outcomes. To capture these multi-level dynamics, we developed a conceptual framework (**Fig. 1A**) specifying hypothesized relationships among patient-, nurse-, and unit-level factors and their influence on documentation practices and patient outcomes. At the <u>patient level</u>, we included comorbidities,^30,31^ smoking status, respiratory function estimates, documentation volume, and patient care activity estimates. Specifically, documentation volume in our analysis refers to shift-based frequencies of nursing flowsheet entries for a specific clinical need, such as vital signs monitoring. Furthermore, to account for individual variance in documentation practice at the <u>nurse level</u>, we identified each patient shift’s primary registered nurse (RN) as the RN contributing the greatest documentation volume or EHR interaction duration for the index patient, an audit log-based proxy for the nurse most responsible for the patient’s documented care^32^ (**Fig. 1B-C**). For the primary RN’s workloads, we included their patient loads and the total workload acuity for the respiratory cohort^33,34^; both have been linked to nursing-sensitive adverse events when suboptimal^35^. At the <u>unit level</u>, we included unit patient volume, Admissions/Discharge/Transfer (ADT) events, and the levels of care (ICUs vs. ACUs).

**Figure 1.**
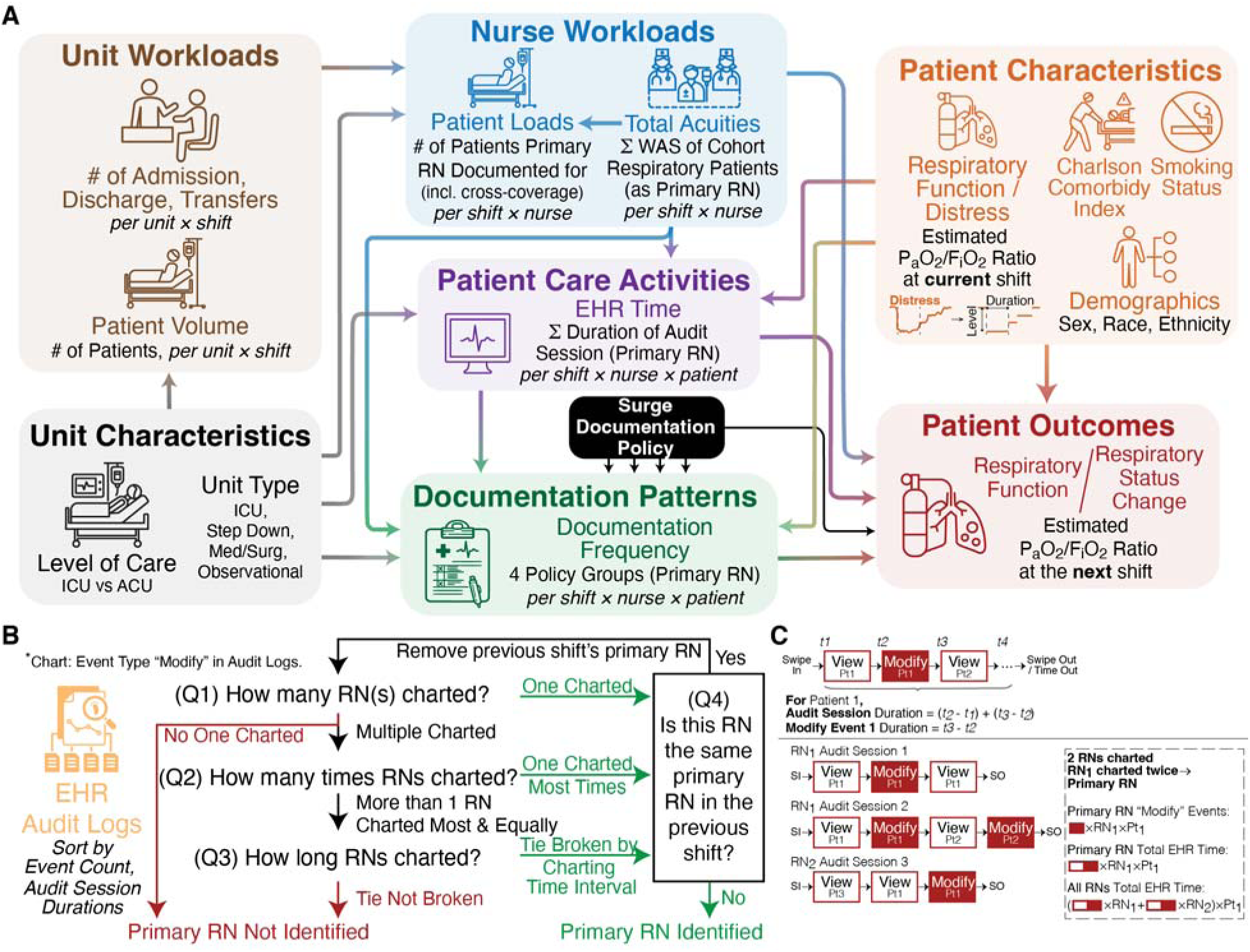
Study Framework and Algorithms to Identify the Primary RN and Calculate EHR Durations from EHR Audit Logs. **A**) the study framework illustrating the key factors and their relationships; **B**) the algorithm we adapted from Riman et al^32^ to identify the primary RN for a certain patient-shift; **C**) the algorithm to calculate the EHR durations (i.e., duration for record-modify (“Modify”) events only, and duration for all logged EHR events [“Modify” events and non-charting EHR events, such as browsing and monitoring]).RN, registered nurse; PCT, patient care technician; ICU, intensive care unit; ACU, acute care unit; Med/Surg, medical and surgical unit; EHR, electronic health records; WAS, workload acuity score (retrospectively calculated using a site-specific formula; see Methods); SI, swipe in; SO, swipe out/time out.

We studied a cohort of adult inpatients with moderate-to-severe respiratory impairment across 54 inpatient units over four years (46 months), spanning pre-, during-, and post-relaxation-policy periods. We applied confirmatory factor analysis (CFA)^36^ to validate latent factor structures, and structural equation modeling (SEM)^37–39^ to examine how these factors influence patient outcomes. Our hypotheses were (1) nurses’ documentation was driven by patient care needs rather than administrative compliance, and (2) reduction of compliance-driven documentation had only a negligible effect on patient outcomes. Our findings have direct implications for future evidence-based reforms to documentation policies and staffing models that aim at reducing documentation burden and freeing nurses’ time and attention for direct patient care.

## RESULTS

Our analyses demonstrate that clinically essential and compliance-driven nursing documentation are empirically separable: when permitted, nurses reduced compliance-driven documentation while maintaining respiratory care documentation, with no impact on patient respiratory outcomes. Below, we describe the cohort and workload profiles across unit types, quantify changes in documentation practices (i.e., durations and volume) in response to the relaxation policy, and present SEM models linking the patient-, nurse-, and unit-level factors to patient respiratory outcomes.

### Cohort Description

Our cohort comprised 520,357 patient shifts from 36,321 unique patients and 44,507 hospital stays across 54 inpatient units between March 1, 2019 and December 31, 2022. Patient demographics and outcome distributions are summarized in **Table 1**. The cohort had slightly more males (**53.94%**) than females and was predominantly white (**81.87%**), with a mean Charlson Comorbidity Index of 0.85 (1.51). Among the 54 units, 9 were ICUs (136,103 shifts, **26.16%**), and 45 were ACUs (384,254 shifts, **73.84%**), with the medical and surgical units (Med/Surg) comprising the largest share (336,757 patient-shifts, **64.72%**).

**Table 1.**
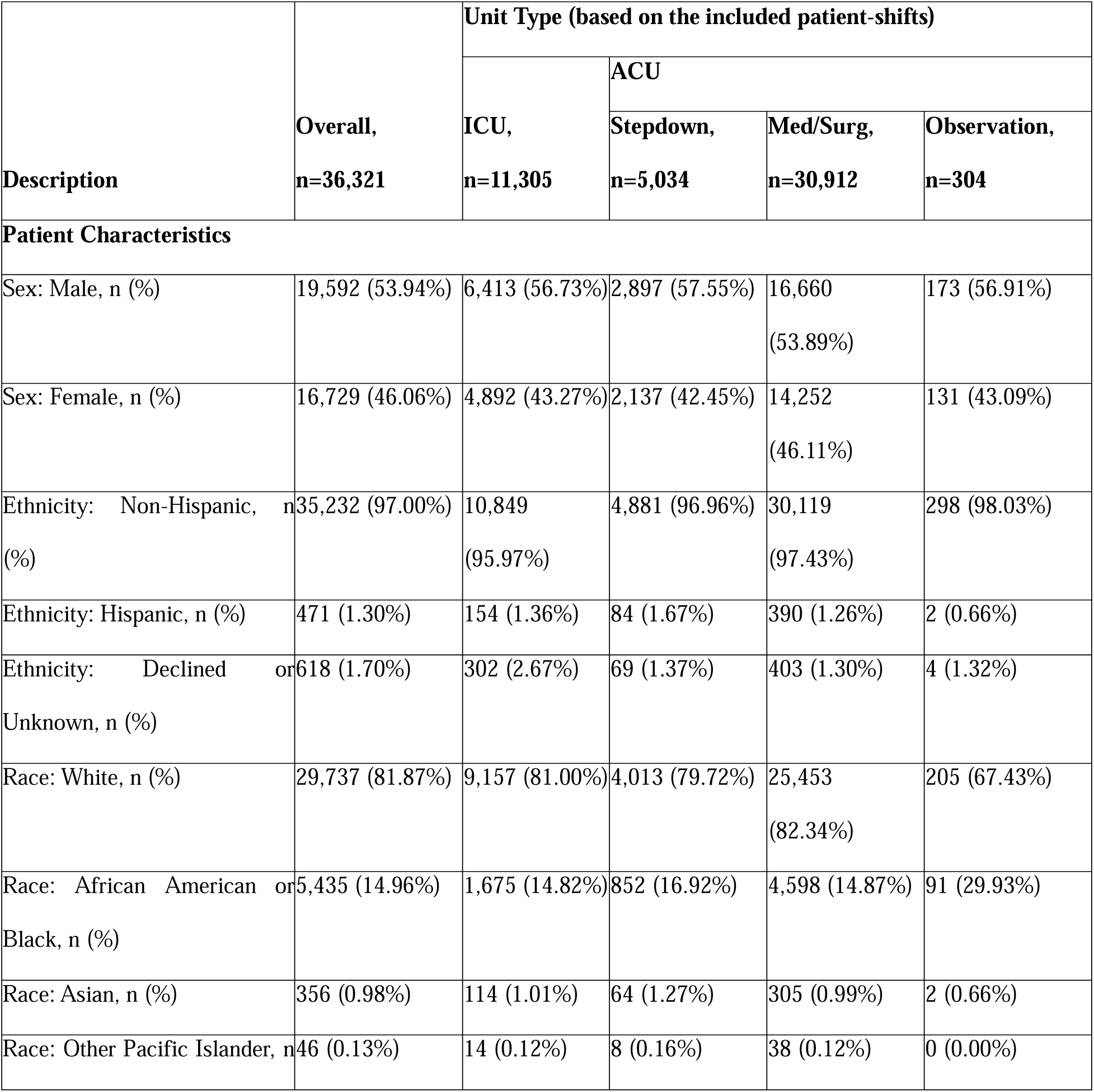
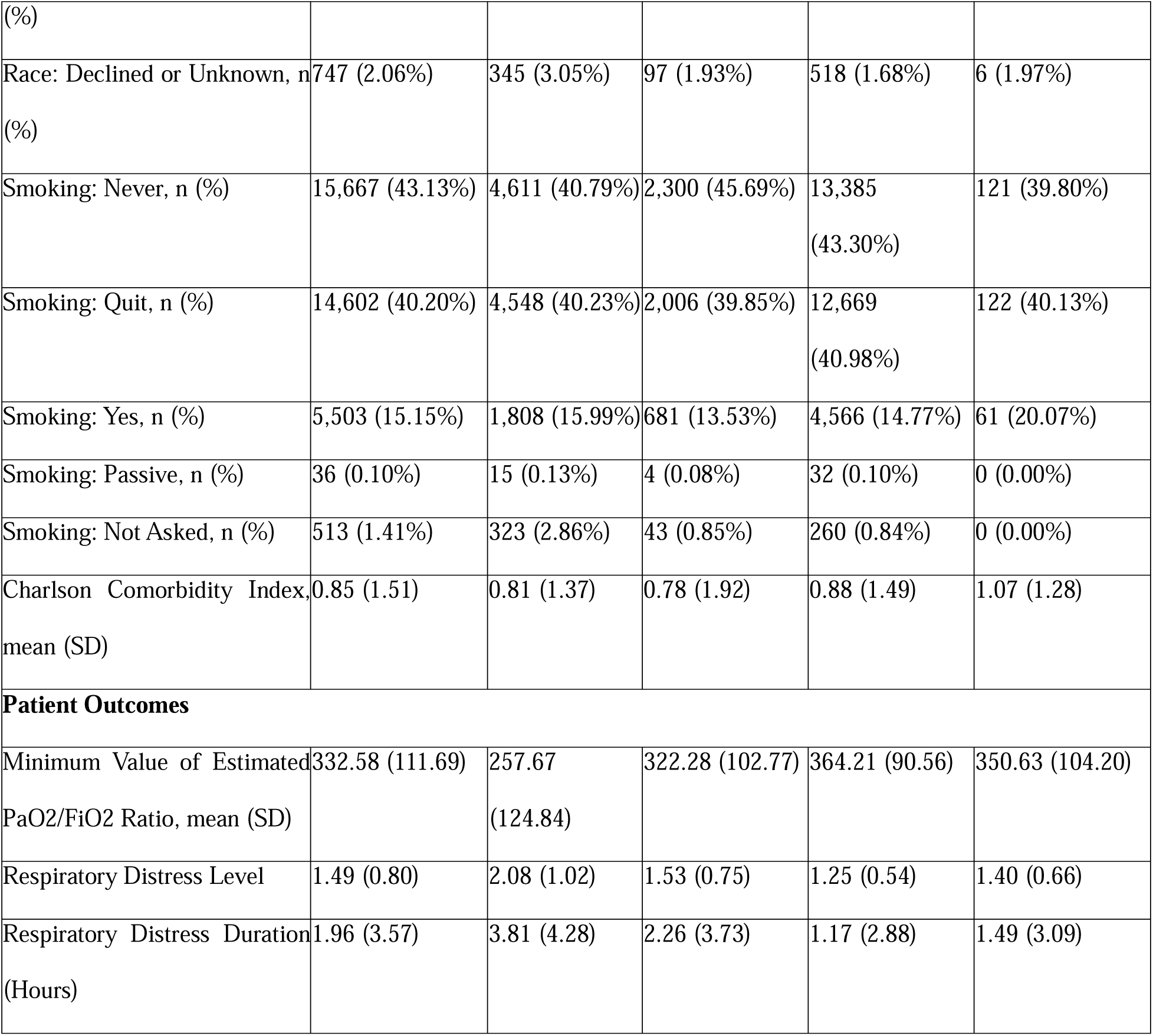
Patient Characteristics and Outcomes. The counts and percentages are based on the number of patients who stayed in the unit for at least one shift. **Respiratory distress level** is an ordinal severity category derived from the minimum estimated P/F ratio per shift (1 = Near Normal Respiratory Function, through 4 = Severe Respiratory Distress), and **respiratory distress duration** is the time the patient remained at their worst distress level during the shift; full definitions are provided in **Methods** and **Supplementary Table S6**. ICU, intensive care units; ACU, acute care units; Med/Surg, Medical and surgical floors; SD, standard deviation; SpO_2_, oxygen saturation; PaO_2_, partial pressure of arterial oxygen; FiO_2_, fraction of inspired oxygen.

The most frequent primary RN identification type was “Multiple nurses charted, but one charted most” (**Fig. 2A**). We used the minimum estimated PaO2/FiO2 (P/F) ratios per shift as a continuous physiological marker of patient respiratory outcomes (**Table 1**, **Fig. 2B-C**). We selected respiratory function as the patient outcome because respiratory care is a core, frequently performed component of nursing practice on both ACUs and ICUs that applies to hospitalized patients broadly while still capturing the COVID-19 population central to the natural experiment. Mean P/F ratio minimums remain relatively stable in Med/Surg and stepdown units, while ICUs showed decreases in late 2020 and early 2022 (**Fig. 2C**); these ICU declines coincided with surges and occurred in the units where the Surge Documentation policy did not apply. Moreover, oxygenation monitoring-related documentation volume, including oxygen therapy details, increased concurrently during these periods (**Fig. 2D**).

**Figure 2.**
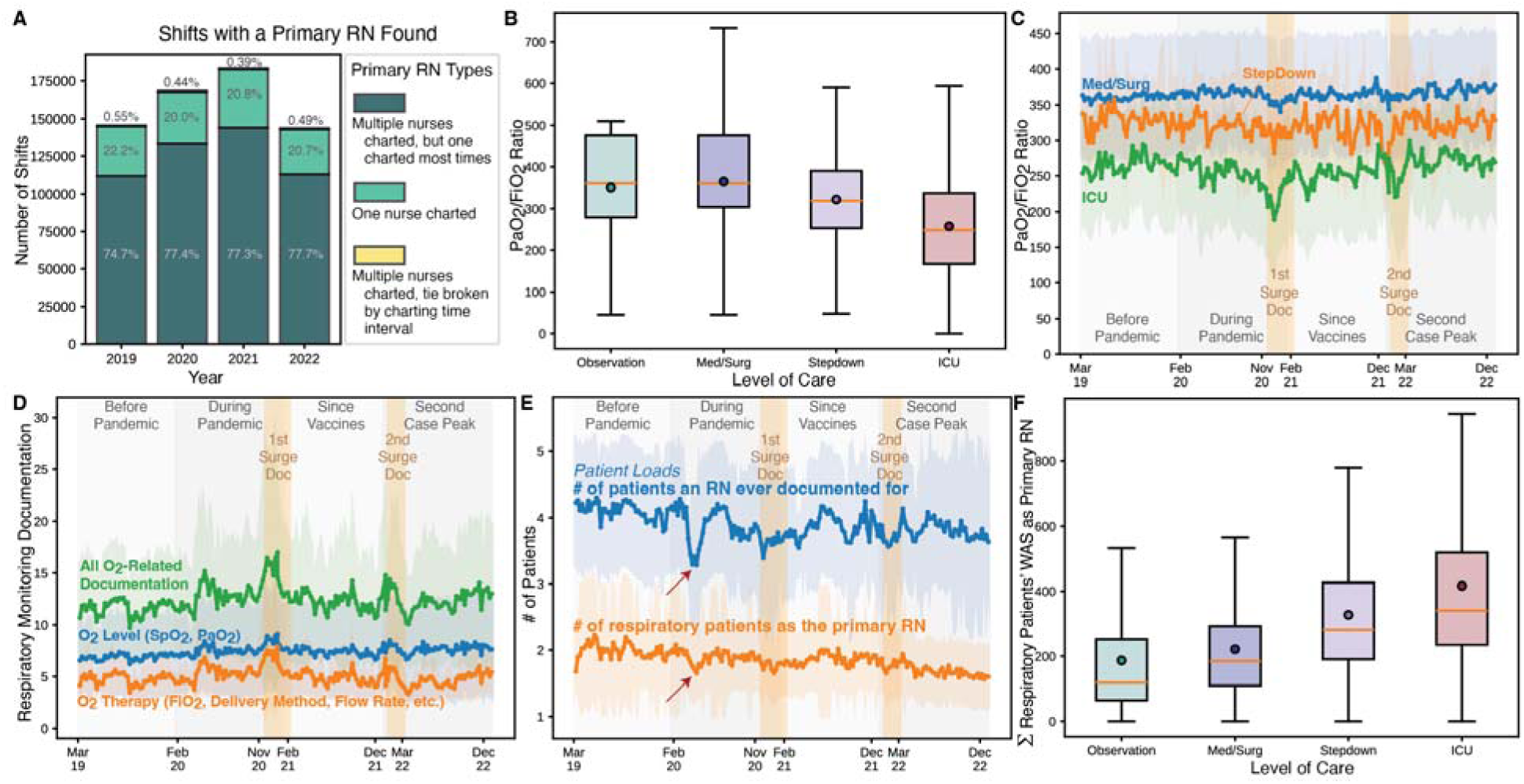
Patient-, Nurse-, and Unit-Level Features. **A**) category distribution and annual percentages when we found a primary RN for a patient-shift successfully; **B**) unit differences of patient outcomes (P/F Ratio minimum); **C**) patient outcome trends across unit types; **D**) temporal trends of oxygenation-related documentation volume across unit types; **E**) temporal trends of patient loads and respiratory patient counts; **F**) unit differences of the primary RN’s total acuity adding up respiratory patients’ WAS. RN, registered nurse; P/F Ratio, PaO_2_/FiO_2_ Ratios; Surge Doc, Surge Documentation policy implementations; Med/Surg, medical and surgical unit; ICUs, intensive care units; WAS, workload acuity score.

### Nurse and Unit Workloads Diverged Sharply Between Critical and Acute Care

Nurse- and unit-level workload profiles differed substantially across levels of care (ACU vs ICU). Nurse-level workloads were estimated using two variables, *total acuities* and *patient loads*. *Total acuities* captured the competing respiratory management needs among our cohort’s respiratory patients assigned to the same primary RN (**Fig. 2F**). The acuities were measured with the Workload Acuity Scores (WAS), which showed comparable annual distributions but significant differences across unit level of care (**Supplementary Fig. S1C-D**). *Patient loads*, a complementary EHR-derived patient-nurse ratio accounting for all patient care needs, was also included (**Fig. 2E**). In ACUs, a nurse generally documented for 4.35 patients per shift and served as the primary RN for 1.99 cohort respiratory patients. In contrast, in ICUs, a nurse took care of 2.65 patients and served as the primary RN for 1.48 cohort respiratory patients on average (**Supplementary Fig. S1E**). Because both measures were derived from EHR activities in audit logs rather than staffing assignments, they are not equivalent to real-world staffing ratios: *patient loads* count every patient a nurse documented for, including brief cross-coverage (e.g., during colleagues’ meals and breaks), whereas primary RN-related counts and acuities include only patients in our respiratory cohort and exclude the remainder of the nurse’s assignment. *Patient loads* dropped sharply in early 2020 in response to the first surge wave of COVID-19 cases (**Fig. 2E**). Because *patient loads* reflect the number of patients each primary RN documented for, this decline indicates fewer patients assigned per nurse during the initial surge, consistent with staffing intensification.

Unit-level workloads were estimated by *ADT events* and *patient volume* on the unit, which were included as covariates to capture shift-level unit activity as they scale with unit size and staffing. Averaged *ADT events* and *patient volume* were comparable across unit types, but acute care units showed a wider distribution with more shifts experiencing high patient turnover (**Supplementary Fig. S1F-G**). Together with the nurse-level workload differences described above, these profiles motivate the multi-group analytic approach used in the SEM models below.

### Rising EHR Time Reflected Non-Charting Activity, Not Active Documentation

Increases in nurses’ EHR time across the pandemic were driven by non-charting EHR activities, such as browsing, reviewing, and monitoring, rather than by active documentation. To establish this, we used total EHR time from audit log analysis as a surrogate measure of *patient care activities* delivered by the primary RN, computed in two ways: for all logged EHR interactions (total EHR time, including read-only, non-charting activities such as chart review and navigation), and for record-modifying events only (i.e., active documentation as labeled “Modify” in audit logs, such as entering or editing flowsheet data and writing notes; hereafter “Modify” events).

**Fig. 3A** shows weekly trends in EHR durations in ACUs for both measures, stratified by primary RN only and by all RNs on the care team. Primary RNs consistently accounted for 60 – 70% of total documentation duration across nurse care teams (“Modify” events: primary RN 0.44 hours vs. all RNs 0.60 hours; all events: primary RN 2.19 hours vs. all RNs 3.55 hours, **Fig. 3A**). A sharp spike in EHR duration occurred across all care settings in early 2020 (ACU trends in **Fig. 3A**, ICU trends in **Supplementary Fig. S1H**). Notably, this spike coincided with the concurrent decline in *patient loads* (**Fig. 2E**), suggesting that despite caring for fewer patients during the initial COVID-19 surge, per-patient care intensity and documentation volume increased substantially.

**Figure 3.**
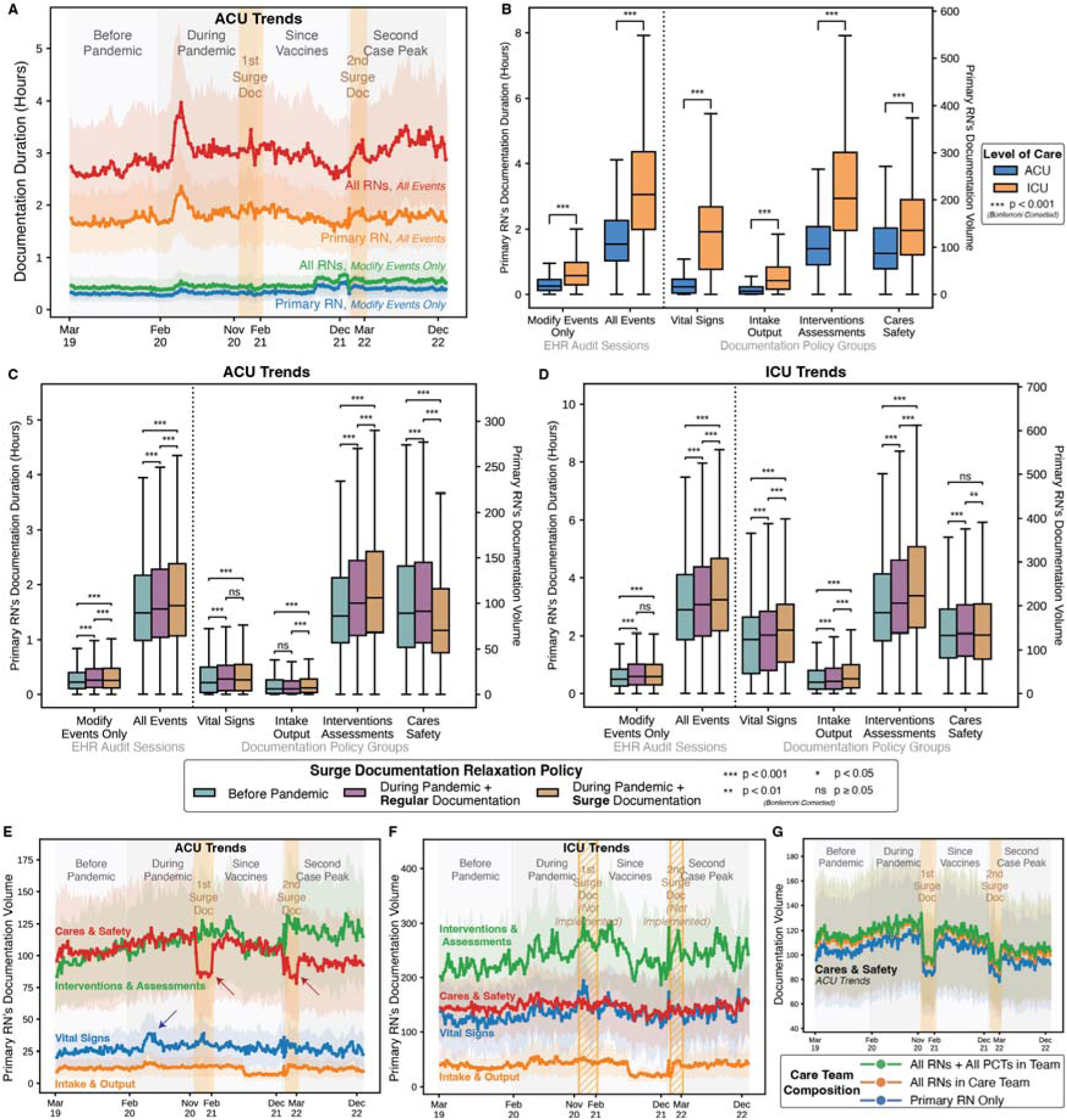
EHR Duration (Patient Care Activity Estimates) and Documentation Volume. **A**) temporal trends for ACU documentation durations; **B**) comparison of documentation duration and volume across levels of care; **C**) comparison of documentation duration and volume across pandemic phases and documentation policy in ACUs; **D**) comparison of ICU documentation duration and volume across pandemic phases and documentation policy; **E**) temporal trends for ACU documentation volume; **F**) temporal trends for ICU documentation volume; **G**) association of Cares & Safety documentation volume with care team composition. Surge Doc, Surge Documentation policy implementations; EHR, electronic health records; RN, registered nurse; PCT, patient care technician; ACU, acute care units; ICU, intensive care units.

EHR durations also differed across levels of care. **Fig. 3B** compares duration effect sizes between ACU and ICU primary RNs. ICU primary RNs spent approximately twice as long on “Modify” events as ACU primary RNs (0.72 hours vs. 0.34 hours, p < 0.001, effect size r = 0.46), with a consistent pattern for all-event durations (3.34 hours vs. 1.78 hours, p < 0.001, r = 0.56). **Fig. 3C-D** shows duration effect sizes within each care setting across policy periods. In ACUs, “Modify” event durations differed significantly across all three periods (before pandemic, regular documentation, and Surge Documentation; **Supplementary Table S2**). In contrast, in ICUs where the Surge Documentation policy was not implemented, “Modify” event durations showed no significant difference between regular and surge documentation periods (p = 1.0, r = −0.01).

Consistent with the divergence stated above, the all-event duration increased progressively across all three periods across unit types, even in ICUs where no relaxation policy was implemented, yet “Modify” event durations displayed a much flatter change between regular and surge periods in both settings (ACU r = −0.02, ICU r = −0.01). Together, these patterns indicate that non-charting EHR activities (all-event duration minus “Modify” events), such as browsing, reviewing, and monitoring, accounted for the increase in duration rather than active documentation (“Modify” events).

### A Hybrid LLM-Expert Approach Profiled Documentation at Scale

Classifying hundreds of flowsheet templates against policy criteria is traditionally a labor- and time-intensive manual task. We previously demonstrated that a semi-automatic strategy, large language models (LLMs) classification paired with targeted expert verification, substantially reduced manual effort in a comparably annotation-intensive task, mapping flowsheet data elements to standard nursing terminology^40^. We adapted this hybrid approach to each policy change in two sequential steps: an *inclusion* step, deciding whether each flowsheet template represented bedside nursing documentation covered by the Surge Documentation policy, and a *policy group assignment* step, assigning each included template to one of the four policy groups (a) Vital Signs (1 policy), (b) Intake/Output (2 policies), (c) Interventions & Assessments (6 policies), and (d) Cares & Safety (20 policies), validated by domain experts (**Supplementary Table S3**). We repeated the query of each template to LLMs 10 times (prompt in **Supplementary Box S1**), and a confidence score was computed as the proportion of affirmative responses; templates scoring above 0.5 were retained. Domain experts verified all policy assignment decisions. Among 727 flowsheet templates, LLM-based classification achieved accuracies of 0.77 in the template inclusion step and 0.94 in the policy group assignment step (**Supplementary Fig. S1I**). The lower accuracy at the inclusion step reflects the inherent ambiguity in distinguishing bedside nursing templates from other specialty templates, where clinical nomenclature overlaps across roles. Because some long templates encompass various patient care needs and relate to multiple policies, we further expanded the classification to the template-measure pairs. LLMs assisted in assigning each of the 5,766 pairs to one of the policy groups three times, with “Not Any Group” (NAG) as an allowable assignment; final decisions were based on the majority vote. A total of 119 pairs with no majority consensus or a NAG majority were subsequently reviewed and manually assigned by domain experts in our research team.

### Nurses Reduced Compliance-Driven Documentation While Maintaining Essential Documentation

*Documentation volume* was analyzed in accordance with the Surge Documentation relaxation policies within four policy groups (Vital Signs, Intake/Output, Interventions & Assessments, and Cares & Safety). As shown in **Fig. 3B**, ICU primary RNs documented significantly more than ACU primary RNs across all four policy groups (all p < 0.001, **Supplementary Table S2**), with the largest differences in Vital Signs (ACU 27.5 vs. ICU 134.7; r = 0.70) and Interventions & Assessments (ACU 110.9 vs. ICU 241.7; r = 0.59). **Fig. 3C-D** shows volume effect sizes within each care setting across policy periods, and **Fig. 3E-F** shows longitudinal volume trends.

In ACUs, primary RNs reduced Cares & Safety documentation in response to policy change, including entries such as fall risk assessments and restraints (before pandemic 103.1, regular documentation 106.4, surge documentation 86.2; regular vs. surge: p < 0.001, r = −0.19). Simultaneously, documentation increased across acuity-related groups, with the largest overall effect in Interventions & Assessments policy group (before pandemic 99.1, regular documentation 113.6, surge documentation 121.1; regular vs. surge: p < 0.001, r = 0.05). Vital Signs volume rose from pre-pandemic to both pandemic periods but showed no significant change between regular and surge documentation periods (p = 0.8951, r < 0.01), indicating a pandemic-driven rather than policy-driven rise.

In contrast, ICUs showed no significant changes in Cares & Safety volume between pre-pandemic and surge documentation period (p = 0.1055, r = 0.02), though acuity-related documentation increased significantly relative to pre-pandemic period across all three groups. Notably, ICU Cares & Safety volume rose during the regular documentation period (p < 0.001, r = 0.04) before returning near pre-pandemic levels during the surge period (p = 0.0027, r = −0.02), suggesting a degree of informal behavioral adjustments in ICUs despite no formal policy mandate. Because the Cares & Safety volume from all RNs and all patient care technicians (PCTs) did not increase to compensate for the primary RN’s volume decrease (**Fig. 3G**), the observed decline reflects a true reduction in volume rather than a redistribution of DocBurden from the primary RN to other nurses or PCTs on the care team.

### The Influence of Documentation Volume and Other Factors on Patient Outcomes: SEM Models

In the following paragraphs, we present four SEM models explaining whether and how documentation volume and other factors influenced patient outcomes. Prior to SEM models, CFA, which tests whether groups of observed variables cohere into the hypothesized underlying constructs, confirmed an excellent measurement model fit for two latent variables, *unit-level workloads* and *documentation* (**Table 2**, **Supplementary Table S4**). Nurse-level workload and patient respiratory distress could not form single latent variables due to opposing factor loading directions and were therefore modeled as individual observed variables. SEM models were estimated on a sub-cohort of 436,248 patient shifts, while 84,109 shifts were dropped because of unavailable next-shift outcomes. Given the large sample size, we prioritized clinical over statistical significance, retaining paths with standardized coefficients over 0.05 as an empirical threshold for clinical effect size (full models in **Supplementary Fig. S2**).

**Table 2.** Model Fit Statistics.

|  |  | CFI<sup>*</sup> | RMSEA<sup>**</sup> | SRMR<sup>***</sup> |
| --- | --- | --- | --- | --- |
| Confirmatory Factor Analysis (CFA) |  | 0.995 | 0.035 | 0.017 |
| Structural Equation Modeling (SEM) | **Model 1** (P/F Ratios as Continuous Measure) | 0.955 | 0.066 | 0.039 |
|  | **Model 2** (P/F Ratios as Ordinal Measure) | 0.963 | 0.052 | 0.027 |
| Multi-Group SEM | **Model 1** | 0.917 | 0.071 | 0.056 |
|  | **Model 2** | 0.921 | 0.058 | 0.040 |

<sup>\*</sup>Comparative Fit Index (CFI) ranges between 0 and 1, with a value greater than 0.90 indicating a good fit and greater than 0.95 indicating an excellent fit.<sup>49</sup>
<sup>\*\*</sup>A Root Mean Square Error of Approximation (RMSEA) below 0.05 indicates excellent fit, and below 0.08 indicates acceptable fit, while an RMSEA of more than 0.1 indicates poor fit.<sup>50</sup>
<sup>\*\*\*</sup>A Standardized Root Mean Square Residual (SRMR) of less than 0.08 is generally accepted as a good fit.<sup>49</sup>

To characterize respiratory outcomes, we applied two complementary models. **Model 1** treated *P/F ratio*s as a continuous physiologic measure (**Table 2**, **Fig. 4A**) so that full quantitative information was preserved. Because clinical decision-making is typically guided by transitions across severity strata rather than small absolute changes in P/F ratio, **Model 2** categorized P/F ratios into ordinal *respiratory distress levels* and estimated directional change in oxygenation over time (*distress duration*). **Model 2** also classified patients as experiencing deterioration, stability, and improvement in respiratory function (*respiratory status change*, **Table 2**, **Fig. 4D**), as patient outcomes. Both models achieved acceptable to excellent model fit (**Table 2**).

**Figure 4.**
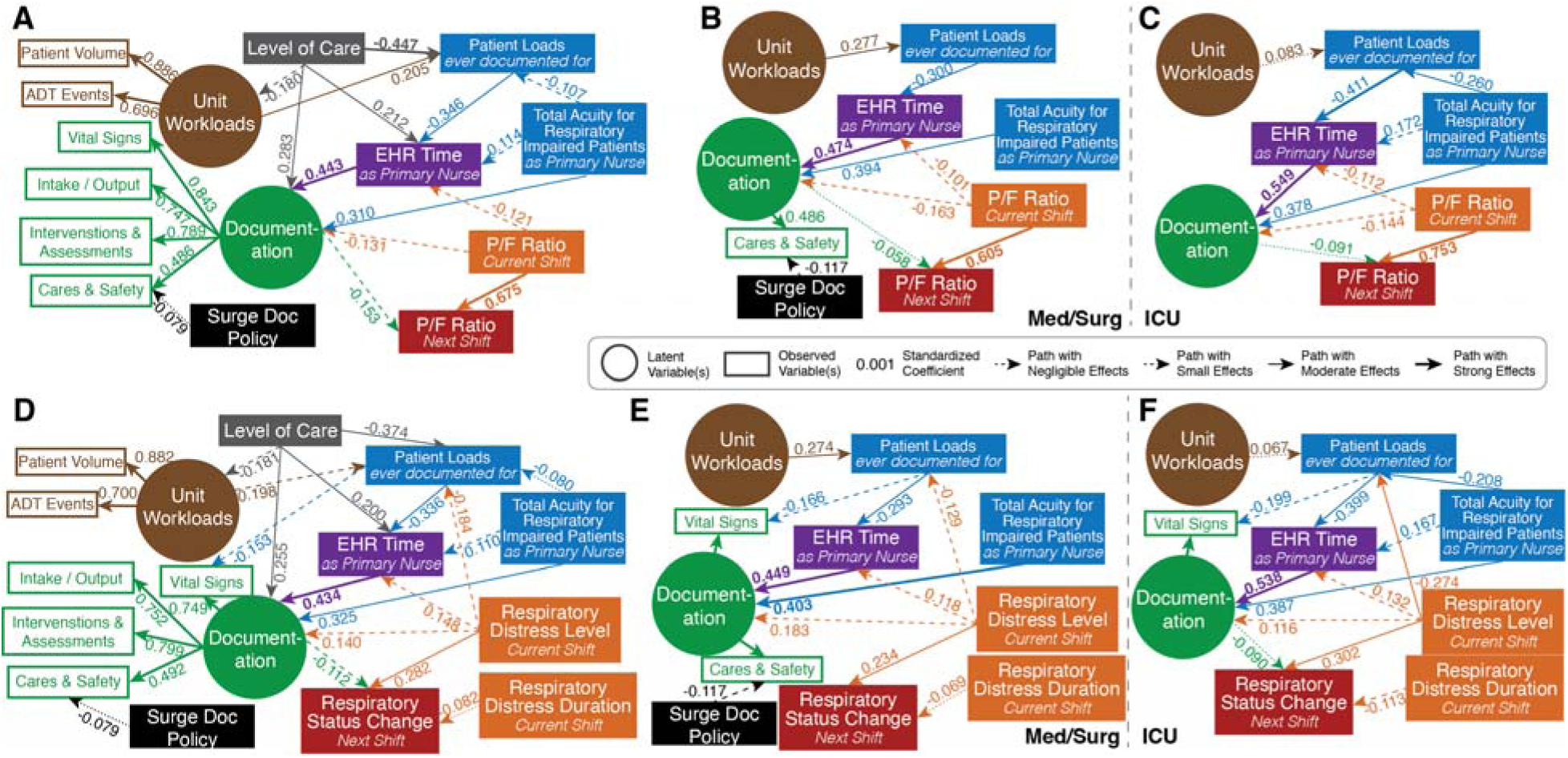
SEM path diagrams with standardized coefficients. **A**) simplified Model 1 path diagram (continuous P/F ratio); **B**) simplified Model 1 multi-group path diagram for Med/Surg; **C**) simplified Model 1 multi-group path diagram for ICUs; **D**) simplified Model 2 path diagram (ordinal respiratory distress, i.e., deterioration based on ΔP/F Ratio); **E**) simplified Model 2 multi-group path diagram for Med/Surg; **F**) simplified Model 2 multi-group path diagram for ICUs. P/F Ratio, PaO2/FiO2 ratio; EHR, electronic health records; ACUs, acute care units; ICUs, intensive care units; Surge Doc, the Surge Documentation policy implementations.

Multi-group SEM analysis was subsequently performed to examine care setting-specific dynamics, stratified by unit types, especially medical/surgical (Med/Surg) units (**Fig. 4B, 4E**) and ICUs (**Fig. 4C, 4F**). These models maintained acceptable fit (**Table 2**), supporting the validity of comparing structural paths across clinical settings. Unless otherwise specified, reported coefficients are from **Model 2**.

### Effects of Surge Documentation Policy: Nurses’ Prioritization of Patient Care over Non-Essential Documentation

The *Surge Documentation policy* was associated with a reduction in *Cares & Safety* documentation volume in ACUs (β = −0.079), with no comparable effect observed in other policy groups (**Supplementary Fig. S2**). Consistent with the temporal trends in **Fig. 3E**, this selective reduction indicates that nurses deprioritized Cares & Safety flowsheets under surge conditions, reflecting a clinical judgment that this category of documentation was relatively non-essential for managing patients’ respiratory status. Critically, this reduction occurred without measurable harm to patient respiratory outcomes, providing direct empirical evidence that targeted reduction of non-essential documentation is achievable without compromising respiratory outcomes examined in this study.

### Current Respiratory Status, Not Documentation, Predicted Future Patient Respiratory Outcomes

Across both modeling strategies, current respiratory function emerged as the dominant predictor of subsequent respiratory status. In **Model 1**, a higher baseline *P/F ratio* was strongly associated with better future respiratory function (β = 0.675; total effects accounting for *EHR time* and *documentation volume:* β = 0.703, **Table 3**). In **Model 2**, current *respiratory distress level* was positively associated with subsequent *respiratory status change* (β = 0.282). Given the ordinal coding scheme (higher *distress levels* = worse baseline; positive change = improvement), this reflects regression towards recovery: patients in more severe distress were more likely to improve over the subsequent shift. *Respiratory distress duration* showed a clinically negligible negative association with improvement (β = −0.082), indicating that prolonged exposure to suboptimal respiratory status slightly reduced the likelihood of short-term recovery. By contrast, *EHR time* per patient showed a clinically negligible direct association with patient respiratory outcomes (β = −0.012 in full model; **Supplementary Fig. S2**). In other words, we found no meaningful direct association between the primary RN’s EHR engagement (as a surrogate of *patient care activities*) and patient respiratory outcomes.

**Table 3.**
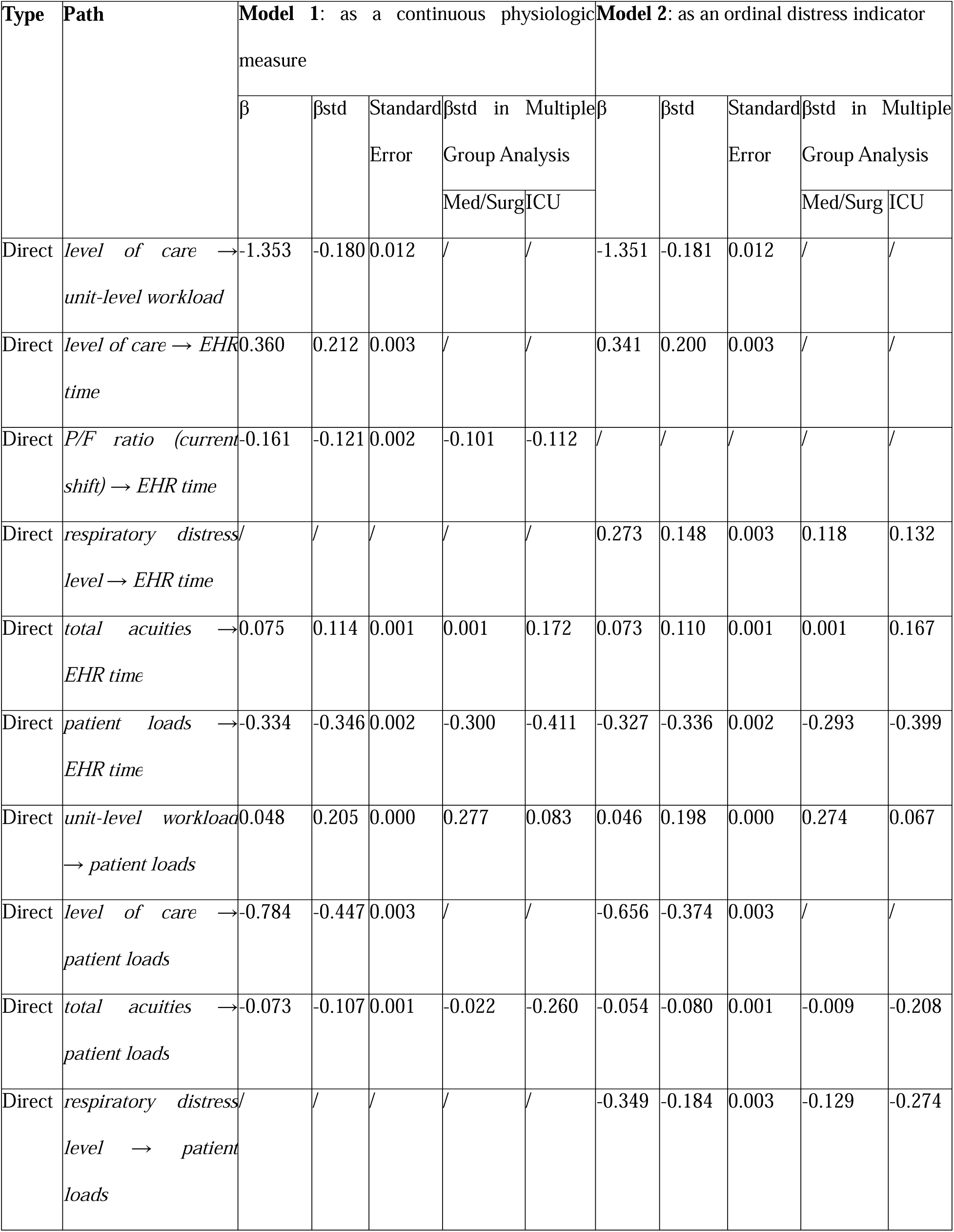
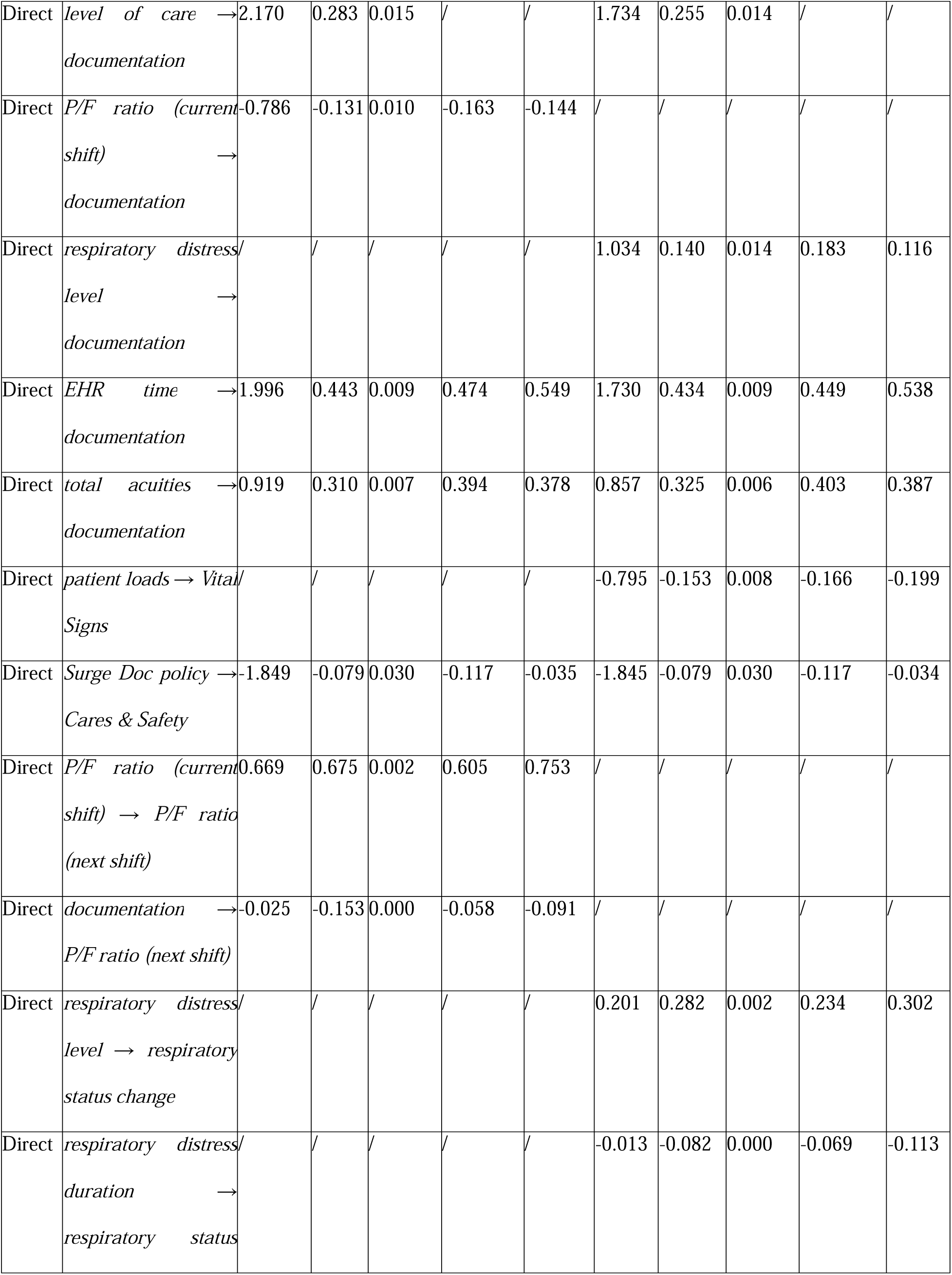
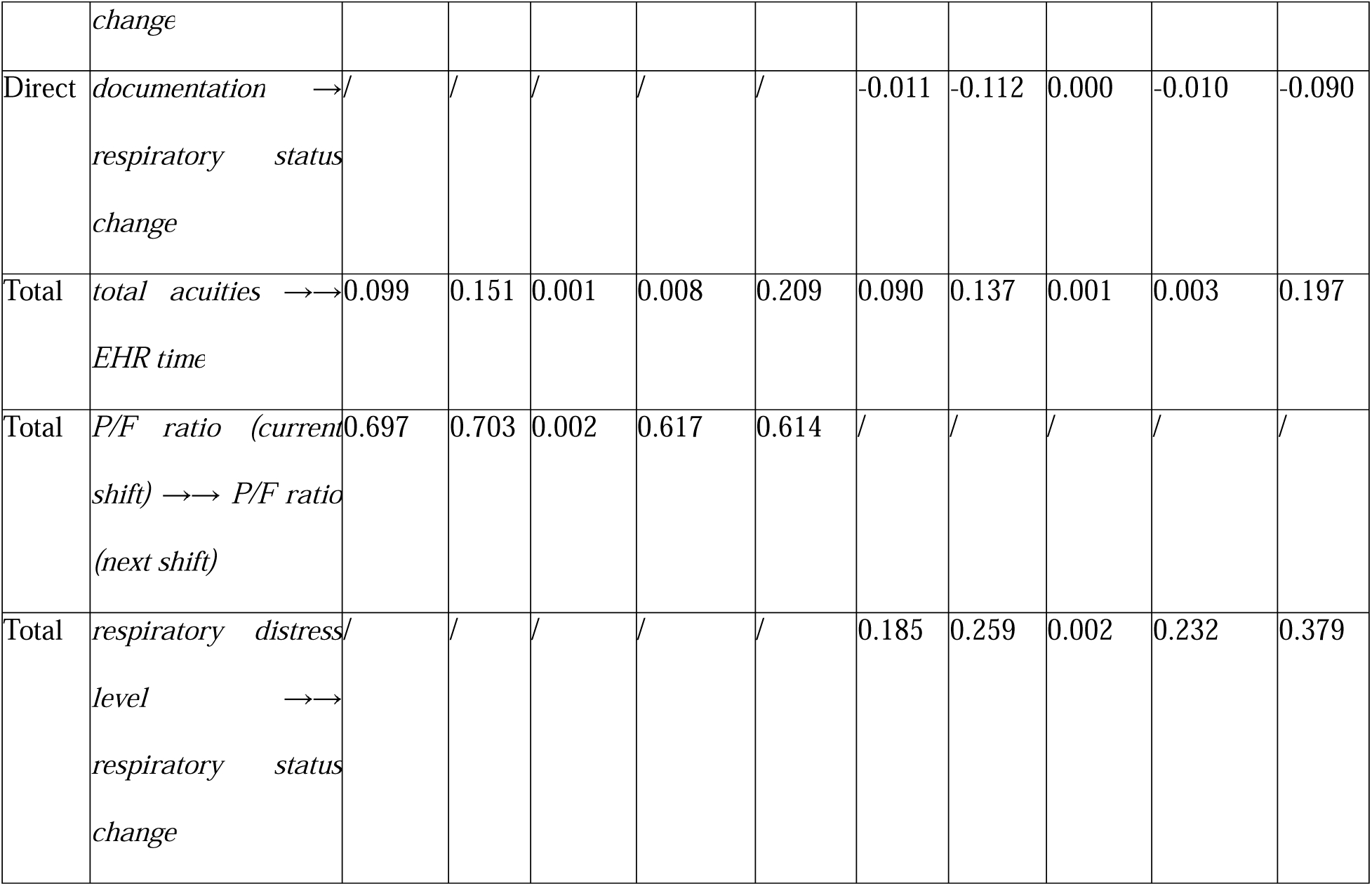
Path models from SEM.

The SEM models also reveal that *documentation* captured concurrent patient status, as more severely ill patients were documented more intensively, as indicated by the negative association between current-shift P/F Ratio and *documentation* (β = −0.131; **Fig. 4A**), and the positive association between current-shift *respiratory distress level* and *documentation* (β = 0.140; **Fig. 4D**). This pattern was consistent across care settings (**Fig. 4B – C, E – F),** confirming that documentation volume tracked real-time clinical status and captured heightened clinical surveillance regardless of level of care. In contrast, the residual direct effect of *Documentation* on next-shift respiratory outcomes (**Model 1**: β = −0.153, **Model 2** β = −0.112) diverged by unit types. In Med/Surg units, this association was clinically negligible or marginal (**Model 1**: β = −0.058, **Model 2** β = −0.010; **Fig. 4B, E**), indicating no meaningful prospective association between documentation volume and outcomes once concurrent respiratory status and workloads were accounted for. In ICUs, the association was larger (**Model 1**: β = −0.091, **Model 2** β = −0.090; **Fig. 4C, F**) but is best interpreted as residual confounding by patient acuity rather than a causal effect: nurses documented more intensively when patients were sicker, and sicker patients were more likely to remain ill or deteriorate in the next shift. This coupling between acuity and documentation was also tighter in critical care, where acuity more strongly shaped nurses’ time allocation (*total acuities* → *EHR time*: ICU β = 0.167 vs. Med/Surg β = 0.001). Together, these findings suggest that real-time documentation volume may function as a complementary signal of nurse-assessed patient instability, with potential utility in early warning systems beyond physiological alarms.^29^

### Nurse-Level Workload Shaped EHR Time and Documentation Volume

Higher *total acuities* were associated with reduced *patient loads* for primary RN (β = −0.080) and increased *EHR time* the primary RN spent on the index patient (β = 0.110). These associations were more pronounced in ICUs (*patient loads* β = −0.208, *EHR time* β = 0.167), and clinically negligible in Med/Surg units (*patient loads* β = −0.009, *EHR time* β = 0.001), consistent with acuity-driven assignments in the critical care.

Independent of acuity, higher *patient loads* were strongly associated with reduced *EHR time* per patient (β = −0.336), reflecting nursing time being distributed across more patients. *EHR time* per patient was in turn strongly associated with *documentation volume* (β = 0.434), completing the structural chain through which nurse-level workloads shaped documentation volume.

### Workload and Documentation Differed by Unit Level of Care

The *unit workloads* latent variable was defined by *patient volume* (standardized factor loading λ = 0.848) and *ADT events* (λ = 0.713). Multi-group analysis revealed distinct workload structures across levels of care. In Med/Surg units, unit-level workloads were overwhelmingly driven by *patient volume* (λ = 0.976), with *ADT event* (λ = 0.637) contributing moderately. In contrast, in ICUs, *ADT events* loaded more strongly (λ = 1.000) than *patient volume* (λ = 0.621), indicating that patient throughput is the dominant driver of unit-level workloads in critical care environments.

A similar divergence was observed for the *documentation* latent variable, defined by four policy groups: *Vital Signs* (λ = 0.817), *Intake/Output* (λ = 0.775), *Interventions & Assessments* (λ = 0.782), and *Cares & Safety* (λ = 0.518). In Med/Surg units, *documentation* loadings were relatively evenly distributed across *Vital Signs* (λ = 0.604), *Intake/Output* (λ = 0.596), and *Interventions & Assessments* (λ = 0.596). In ICUs, *documentation* was more heavily weighted toward *Interventions & Assessments* (λ = 0.790), reflecting a greater emphasis on therapeutic activities in critical care (**Supplementary Table S5**).

## DISCUSSION

Nursing documentation has long been treated as an undifferentiated obligation; our findings disrupt that assumption. Using a COVID-19 documentation relaxation policy as a natural experiment across 520,357 patient shifts, we provide the first large-scale empirical evidence that clinically essential and compliance-driven nursing documentation are separable: when permitted, front-line nurses reliably distinguished between them. In ACUs, primary nurses reduced Cares & Safety flowsheet entries by 19% (106.4 to 86.2 entries; r = −0.19) while maintaining or increasing documentation tied directly to respiratory care, with no impact on patient respiratory outcomes; the documentation that remained was the documentation that mattered.

One dimension of this evidence is the volume of documentation itself. Documentation volume co-varied with patient instability, suggesting it also encodes a latent clinical surveillance signal; in fact, this clinical surveillance has been demonstrated before in the CONCERN Early Warning System Study^29^. By contrast, the residual association between documentation volume and subsequent respiratory outcomes, which was larger in ICUs than in Med/Surg units, is best interpreted as residual confounding by patient acuity rather than that documentation volume itself altered outcomes. In critical care, patient acuity more strongly shaped nurses’ time allocation and documentation intensity, making it more likely that unmeasured severity manifests jointly in documentation volume and short-term patient trajectories. This tighter acuity-documentation coupling may be reinforced by ICU nurses’ greater autonomy to intensify charting in accordance with their clinical judgment, whereas ACU nurses operating under higher patient loads and more standardized documentation requirements may be less able to deviate from baseline charting patterns. This interpretation is consistent with prior work demonstrating that nurses who voluntarily intensify monitoring and documentation typically do so in anticipation of deterioration rather than in response to administrative prompts, underscoring that nursing documentation volume can reflect clinical knowledge and judgment about patient instability rather than mere record-keeping^29^.

Beyond documentation volume, our results highlight that documentation’s clinical value also depends on *what* is documented, not just how much. Under resource constraints, nurses did not reduce documentation uniformly: they deprioritized Cares & Safety flowsheets, while maintaining or increasing documentation in domains directly relevant to a patient’s respiratory care. This documentation prioritization^41^ pattern supports our core hypothesis that essential documentation is driven by patient care needs rather than administrative compliance, and it is reinforced by two studies from our group on nurses’ own perspectives^41,42^. In a survey of inpatient nurses, 88% reported that at least some required documentation was completed primarily for compliance rather than to support patient care. Documenting the hourly safety checks (“nursing rounds”, part of Cares & Safety documentation) was among the entries most frequently cited as redundant to record; the burden lay in logging that the round occurred, not in performing the check itself, which remains an important part of safe care.^42^ Complementary interviews highlighted the “focused charting” strategy, in which nurses prioritize the data most relevant to a patient’s current condition.^41^ Most importantly, the reduction in Cares & Safety documentation was not associated with measurable harm to patient respiratory function, providing reassurance that reducing compliance-driven documentation need not compromise care. It also speaks to the broader point: nurses do more than documentation can capture. Much of nursing practice occurs through direct assessment, clinical judgment, and timely response to patient needs, which are activities not fully captured and reflected in EHR. Reduced documentation, in other words, did not mean reduced care. Consistent with this, our prior work found that nurses placed little value on templated care plan format (one type of Cares & Safety flowsheets), reporting that “*We did not use them and nothing changed in terms of patient outcomes*”^41^; what they devalued was the rigid template, not care planning itself, which nurses enact through ongoing assessments and responsive documentation. Taken together, these findings suggest that the path to sustainable documentation reform depends not on uniform reduction but on accurately distinguishing which documentation categories are clinically essential versus compliance-driven.

Beyond which categories were prioritized, the Surge Documentation policy also permitted a change in the form of documentation: some Cares & Safety entries were allowed to be recorded in an end-of-shift summary note instead of structured flowsheets. Structured flowsheets entries capture discrete observations at fixed time points, whereas narrative notes integrate clinical context, temporal patterns, and nursing reasoning, thereby encoding clinical knowledge rather than raw data. Nurses themselves recognized this distinction. In our previous study, they reported greater appreciation for the end-of-shift narrative summaries over structured, checklist-style care plan entries.^42^ Although note volume or content were not directly analyzed here, primary RNs maintained comparable EHR “Modify” event durations across policy periods (**Fig. 3C**), a pattern consistent with nursing time being redistributed across documentation types rather than eliminated^41,42^. Taken together, these patterns point to a central implication. Active documentation time remained stable across policy periods even as compliance-driven flowsheet volume declined, and EHR time showed no meaningful direct association with respiratory outcomes. Documentation’s clinical value therefore appears to lie not in the time spent or the volume produced, but in its content and intent: whether it captures nurses’ real-time clinical judgment.

In our study, we used a LLM to assist with documentation profiling, which substantially reduced the manual effort that was traditionally labor- and time-intensive. Classifying 727 flowsheet templates, each queried 10 times, required 62 minutes of LLM runtime and approximately 90 minutes of expert review. Classifying 5,766 template-measure pairs, each queried three times, required 55 minutes of LLM runtime, with only 119 pairs (2% of all combinations) requiring manual adjudication, completed in under 30 minutes. This hybrid approach, combining LLM capabilities with targeted expert verification, enables scalable documentation classification at minimal expert burden. This builds on our prior work applying LLMs to concept mapping from flowsheet data elements to standard nursing terminology^40^, in which a similar semi-automatic strategy reduced manual efforts of a comparably labor-intensive informatics task. These applications of LLMs, along with reports from other literature^43–45^, suggest that this strategy can generalize to other annotation- and classification-intensive tasks in clinical informatics, offering substantial savings in time and expert resources where exhaustive manual review will otherwise be required.

We acknowledge several limitations. First, we used a site-specific formula to calculate the workload acuity scores. Although it ensures consistency in calculations, this approach may under- or over-estimate nursing workloads when flowsheets are retired or newly adopted. These scores were also not used by hospital administration to guide staffing decisions during the study periods, limiting their validity as proxies for actual workload allocation. Future work should incorporate operational staffing metrics to estimate nursing workloads. Second, a formal washout period for policy transitions was not included since several educational materials on the Surge Documentation policies were handed out before the policy implementation. Furthermore, we observed a rapid decrease in Cares & Safety documentation volume, suggesting a rapid behavioral adaptation by bedside nurses to the policies. Third, unobserved unit-specific policies may have influenced documentation patterns and EHR time estimates. Future prospective studies with more granular policy documentation can reduce such effects to a minimal extent and isolate practice variation. Fourth, our outcome was scoped specifically to respiratory status, as it captured but was not limited to the COVID-19 population central to our natural experiment. However, some Cares & Safety documentation supports other nursing-sensitive outcomes, such as falls and pressure injuries, which we did not assess. Whether reduced documentation of these activities affects those outcomes remains an important question for future study. Finally, this study was conducted at a single Midwestern academic medical center in the US with a specific EHR infrastructure. As staffing models and documentation cultures vary across institution types, the generalizability of these findings to other community hospitals or systems with different nurse-to-patient ratios or EHR vendors remains to be established. Multi-site replication, particularly in settings with greater patient diversity and resource constraints, is an important direction for future work.

Taken together, our findings carry direct implications for documentation reform and policy. Rather than reducing documentation indiscriminately, health systems should target compliance-driven categories as the primary site of burden relief, preserving the clinical essential documentation that front-line nurses already prioritized when given the opportunity to do so. These results lay an empirical foundation for policies that align documentation requirements with clinical value: to reduce nurse burden and to preserve the integrity of patient care, simultaneously. Cut compliance, not autonomy.

## METHODS

### Study Design and Data Source

We conducted a retrospective observational study examining the associations between nursing EHR documentation and patient outcomes related to respiratory care management at a Midwestern academic medical center between March 1, 2019, and December 31, 2022. The study period encompassed temporary documentation relaxation policies implemented during COVID-19 pandemic surge periods, providing a unique natural experiment for evaluating how policy-driven changes in documentation requirements influenced EHR burden and subsequent patient outcomes.

We extracted patient data, nursing documentation, and the linked EHR audit logs from the institution’s EHR vendor database through an internal data brokerage process. The study protocol and all data use were approved by the Institutional Review Board (IRB #202205146).

### Study Population

The patient cohort was derived from the ENDBurden study and included all adult inpatients with moderate-to-severe respiratory impairment. A patient was classified as having moderate-to-severe respiratory impairment if any of the following criteria were met: oxygen saturation less than 94%, a respiratory system-related admission diagnosis, or receipt of at least one sedation medication during an ICU stay. Using physiological criteria, rather than relying solely on diagnosis codes, allowed for enhanced stratification of patient acuity for outcome comparisons. We excluded the patients with Do Not Resuscitate/Do Not Intubate orders, those receiving hospice and palliative care, and those admitted to pediatric or obstetrics/gynecology units, as their care processes and documentation practices differ from typical respiratory care workflows.

The initial cohort included 38,090 unique patients, with 46,909 unique hospital stays across 56 units. Mean length of stay (LOS) was 11 days (Median: 5.6 days; **Supplementary Fig. S1A**). Stays were expanded into 653,276 shifts (day shift: 7:00:00 AM – 6:59:59 PM, night shift: 7:00:00 PM – 6:59:59 AM^+1^) for shift-based analysis. After excluding shifts without an identified primary RN or with missing values, the final cohort included 520,357 shifts (**79.65%**) from 36,321 unique patients, 44,507 hospital stays, and 54 units.

Each patient’s hospital stay was expanded into shift-level observations (day shifts: 7:00:00 to 18:59:59; night shifts: 19:00:00 to 6:59:59). The following data preprocessing and feature extraction were guided by a conceptual model (**Fig. 1A**) specifying hypothesized relationships among patient-, nurse-, and unit-level factors. When multiple values were available for a given feature within a shift, the median was used. Shifts with missing data in any extracted feature required for analysis were excluded from the final modeling cohort. Data preprocessing, feature extraction, and visualizations were conducted using NumPy-2.0.1, Pandas-2.3.1, matplotlib-3.10.0, and seaborn−0.13.2 in Python 3.12.

### Primary Registered Nurse (RN) Identification

We identified the primary Registered Nurse (RN) for each index patient shift using credentials, provider types, and role classifications documented in EHR (PROV_TYPE is “Registered Nurse” OR ((PROV_TYPE is NULL) AND (CLINICIAN_TITLE contains “RN”))). We then applied a modified version of the algorithm by Riman et al^32^ to select the nurse who contributed the greatest documentation volume or EHR interaction time for the index patient shift (**Fig. 1B, Supplementary Fig. S1B**). Multiple nurses commonly document on the same patient within a single shift, typically reflecting routine coverage during meals and rest breaks, shift-overlap handoffs, and team-based tasks; the primary RN designation identifies the predominant documenter rather than the formally assigned nurse, as staffing assignment records were not available.

### Unit-, Nurse-, and Patient-Level Feature Extraction

**Unit-level features** described the care environment. Unit characteristics included *level of care* (ICU, stepdown, medical/surgical, and observation units) to capture practice variations. *Unit workloads* comprised ADT events and unit patient volume per shift.

To capture **nurse-level features**, we determined the primary RN’s *patient loads* by counting all patients the nurse documented for, regardless of their role within individual care teams.

*Total acuities* were quantified by the Workload Acuity Score (WAS). Because this score was not routinely generated during the study period, it was retrospectively calculated using a site-specific EHR formula. The WAS consists of nine subscales: (i) Activities of Daily Living (ADLs), (ii) Admissions, (iii) Assessments, (iv) Discharges, (v) Lines/Drains/Airways (LDAs), (vi) Orders, (vii) Risks, (viii) Medications, and (ix) Wounds. The subscales (ii) Admissions and (iv) Discharges were excluded from our study because we focused on the influence of shift-based documentation and had included *ADT events* as a unit-level feature. To capture the total bedside needs for respiratory care management, we utilized documentation from all clinicians for calculation, not necessarily restricted to the primary RN. Scores were computed hourly, averaged per shift, and then summed for all cohort respiratory patients assigned to the primary RN.

**Patient-level features** included *patient characteristics*, *patient care activities, documentation patterns*, and *patient respiratory outcomes*. *Patient characteristics* comprised demographics, smoking status, and the Charlson Comorbidity Index (CCI) calculated from the active problem list during the shift.

### EHR Duration Calculation as Surrogate for Patient Care Activities

*Patient care activities* were estimated using the total EHR time the primary RN spent during the index patient shift, derived from the sum of audit session durations in the EHR audit logs (**Fig. 1C**). Audit log events were classified by access type: “Modify” events comprised actions that altered the patient record, e.g., creating or editing flowsheet entries or notes, whereas all-event durations additionally included read-only interactions (e.g., chart review, results viewing, Patient Story displays) and administrative workflows (e.g., printing). Total EHR time was computed across all events; active documentation time was computed across “Modify” events only.

### Hybrid LLM-Expert Approach for Documentation Classification and Volume Calculation

*Documentation volume* was measured as the shift-level flowsheet entry frequency for the primary RN across four policy groups according to the documentation relaxation policy specifics^28^: 1) Vital Signs, 2) Intake/Output, 3) Interventions & Assessments, and 4) Cares & Safety (**Supplementary Table S3**). We used an enterprise version of OpenAI endpoints (gpt-4.1-mini) to filter policy-related flowsheet templates and assign them to a specific policy (prompt in **Supplementary Box S1**). Each flowsheet template was tested 10 times to exclusively identify relevant policies. Flowsheet templates unrelated to bedside nursing workflows were excluded using keyword searches (e.g., emergency department (“ED”), operating room (“OR”, “OP”), physical therapy (“PT”), occupational therapy (“OT”), respiratory therapist (“RT”), rehabilitation (“RHB”), ambulatory medicine (“AMB”), oncology (“ONC”, “CHEMO”), obstetrics/gynecology (“OB”), pediatrics (“PEDS”), etc.). Flowsheet templates not assigned by the LLM, but with more than 500 entries across 4 years, were manually reviewed. Inclusion and exclusion decisions were reconciled with expert validation.

Because some flowsheet templates were assigned to multiple policy groups, such as “ICU Vital Signs and Intake/Output”, we further expanded them with the subordinate measures and populated them into template-measure pairs^40^. The LLM was then used to assign template-measure pairs to a policy group for accurate volume estimates (prompt in **Supplementary Box S2**). Each query provided three consecutive pairs to the LLM to provide context. Therefore, each pair was queried three times. Assignments were determined by majority vote; pairs with three differing assignments were manually adjudicated by domain experts. A random 3% sample of pairs was independently reviewed to validate LLM assignments against expert opinions.

### Statistical Analysis

Group differences displayed in box plots were assessed nonparametrically given the right-skewed distributions of EHR durations and documentation volume. For comparisons between levels of care (**Fig. 3B**), ACU and ICU primary RNs were treated as two independent groups and compared using a two-sided Mann-Whitney U test per measure. To control the family-wise error rate, Bonferroni correction was applied by multiplying each raw p-value by the number of measures tested (k = 6). For comparison within the same level of care but across documentation policy periods (**Fig. 3C – D**), a Kruskal-Wallis test was first applied to test across three periods (before pandemic, during pandemic with regular documentation policy, and during pandemic with Surge Documentation policy); pairwise Mann-Whitney U tests were conducted only when the Kruskal-Wallis test was significant (α = 0.05). All Kruskal-Wallis tests were significant (ACU: all H > 259, all p < 0.001; ICU: all H > 83, all p < 0.001), and pairwise comparisons were conducted for all measures in both settings. Bonferroni correction was applied across all pairwise comparisons (3 pairs * 6 measures = 18). Effect sizes were quantified using the rank-biserial correlation coefficient r, where |r| ≥ 0.10, 0.30, and 0.50 are interpreted as small, medium, and large effects, respectively. All tests were two-sided with a Bonferroni-corrected significance threshold of α = 0.05. All nonparametric tests were conducted using SciPy-1.16.2 in Python 3.12.

### Patient Respiratory Outcome Derivation

*Patient respiratory outcomes* were measured using the *PaO2/FiO2 (P/F) ratio*. Because arterial blood gas analysis was not routinely performed, PaO2 was estimated from peripheral oxygen saturation (SpO_2_) using the nonlinear Ellis equation^46–48^. Only SpO_2_ values less than 97% were used for the imputation of P/F ratios^46^, as values above 97% approach the plateau of the hemoglobin-oxygen dissociation curve, rendering imputation unreliable^46^. Therefore, for those values exceeding 97%, the P/F ratio was capped at 476, representing a non-ventilated patient breathing room air with satisfactory respiratory function.

Two modeling approaches were used to characterize patient respiratory outcomes. **Model 1** treated the *P/F ratio* as a continuous variable, while **Model 2** transformed the continuous P/F ratios into ordinal *respiratory distress levels* **(Supplementary Table S6).** In **Model 2**, we further quantified outcomes by calculating the *distress duration*, defined as the total time the index patient remained at their worst observed respiratory distress level during the shift (**Fig. 1A**). If a patient remained at distress level 1 for the entire shift, the duration was recorded as 0. Within this framework, a superior respiratory outcome was characterized by a lower *distress level* and a shorter *distress duration*.

### Structural Equation Modeling (SEM) Analysis

We employed structural equation modeling (SEM)^37–39^ to quantify relationships among patient, nurse, and unit factors and their association with patient outcomes. Categorical features (e.g., race in *patient demographics*, level of care in *unit characteristics*) were modeled as independent exogenous features, while binary and ordinal features with more than four categories (e.g., smoking status in *patient characteristics*) were treated as continuous. Latent variables were tested using confirmatory factor analysis (CFA)^36^ to test if data fit the theoretical measurement model and to assess model fit. Model fits were evaluated using the comparative fit index (CFI)^49^, the root mean square error of approximation (RMSEA)^50^, and the standardized root mean square residual (SRMR). The CFA and SEM models were estimated using the maximum likelihood estimation with robust (Huber-White) standard errors and a scaled test statistic that is asymptotically equal to the Yuan-Bentler test statistic, implemented in lavaan−0.6-20 in R-4.4.0.

## AUTHOR CONTRIBUTIONS

P.Y., S.R., and H.F. conceptualized and designed the study. H.F. and P.Y. developed the methodology. H.F. curated and managed the dataset. H.F. conducted the data analysis under the supervision and project oversight of P.Y. H.J. served as the statistician and oversaw the statistical analysis. R.M., J.T., and S.R. served as domain experts and verified the findings. H.F. and R.M. drafted the manuscript, with revisions and critical input from P.Y. and S.R. All authors reviewed and approved the final manuscript.

## ADDITIONAL INFORMATION

We used an enterprise version of OpenAI endpoints (gpt-4.1-mini) between October and December 2025 to assist with flowsheet groupings and figure aesthetics. Details are provided in Methods and Supplementary Boxes. Claude Sonnet 4.6 was used for manuscript revision.

## Data Availability

All data produced in the present study cannot be shared due to compliance with the Health Insurance Portability and Accountability Act (HIPAA) requirements.

